# Plasma P-selectin is an early marker of thromboembolism in COVID-19

**DOI:** 10.1101/2021.07.10.21260293

**Authors:** Bánk G. Fenyves, Arnav Mehta, Kyle R. Kays, MGH COVID-19 Collection & Processing Team, Marcia B. Goldberg, Nir Hacohen, Michael R. Filbin

## Abstract

Coagulopathy and thromboembolism are known complications of SARS-CoV-2 infection. The mechanisms of COVID-19-associated hematologic complications involve endothelial cell and platelet dysfunction and have been intensively studied. We leveraged a prospectively collected acute COVID-19 biorepository to study the association of plasma levels of a comprehensive list of coagulation proteins with the occurrence of venous thromboembolic events (VTE). We included in our analysis 305 subjects with confirmed SARS-CoV-2 infection who presented to an urban Emergency Department with acute respiratory distress during the first COVID-19 surge in 2020; 13 (4.2%) were subsequently diagnosed with venous thromboembolism during hospitalization. Serial samples were obtained and assays were performed on two highly-multiplexed proteomic platforms. Nine coagulation proteins were differentially expressed in patients with thromboembolic events. P-selectin, a cell adhesion molecule on the surface of activated endothelial cells, displayed the strongest association with the diagnosis of VTE, independent of disease severity (p=0.0025). This supports the importance of endothelial activation in the mechanistic pathway of venous thromboembolism in COVID-19. P-selectin together with D-dimer upon hospital presentation provided better discriminative ability for VTE diagnosis than D-dimer alone.

## Introduction

Studies have shown that COVID-19 is often complicated by coagulopathy and carries an increased risk of intravascular thrombosis (Helms et al., 2020). Extensive microvascular thrombosis in the lung, kidney, and heart tissue with neutrophil extracellular trap (NET)-formation, which may play a significant role in acute lung injury and the development of acute respiratory distress syndrome (ARDS), has also been reported (Nicolai et al., 2020). The pathomechanism involves the interplay of the coronavirus with endothelial cells, platelets, and immune cells (Iba et al., 2021). The procoagulant imbalance and subsequent thrombotic events are proportional to disease severity and are associated with adverse outcomes (Leentjens et al., 2021). Plasma coagulation proteins that have been associated with intubation status, thrombosis and death, include increased levels of D-dimer, factors V and VIII, von Willebrand factor (vWF), and fibrinogen, as well as decreased levels of ADAMTS13, a vWF-cleaving protease (Goshua et al., 2020; Martín-Rojas et al., 2020; Stefely et al., 2020). However, previous studies investigated only small numbers of coagulation proteins together, and they were limited in their ability to adjust for confounders.

The aim of this post-hoc analysis of a previously published dataset (Filbin et al., 2021) was to explore the association of the plasma levels of 31 coagulation proteins with thromboembolic complications of COVID-19. Plasma protein levels of enrolled patients upon hospital arrival were assayed using two independent proteomic platforms (Olink Explore 1536 and Somalogic SomaScan). We also explored the temporal dynamics of relevant coagulation proteins and study their potential utility as early biomarkers of thromboembolic complications.

## Results

### Clinical parameters - conventional lab parameters

We prospectively enrolled 384 patients presenting to the Emergency Department with respiratory distress. 306 patients were confirmed SARS-CoV-2 positive by PCR, while 78 patients were COVID-19-negative (Table 1). The COVID-positive and negative groups were similar in gender and outcome (illness severity), but the COVID-19-positive cohort was markedly younger and consisted of proportionally more Hispanic patients. Amongst the 306 COVID-positive patients enrolled, 13 (4.2%, 95% CI 2.5-7.1) developed PE or DVT, confirmed by computed tomography with pulmonary-phase angiography (CTPE) or venous duplex ultrasonography (Table 1). Interestingly, incidence of VTE was similar in the COVID-19-negative cohort. COVID-19 patients with VTE were older (median age: 69 (IQR 62, 78) vs 58 (IQR 45, 75), p = 0.2) and had significantly higher rates of severe illness (28-day Acuity max scores of 1 or 2) (85% vs 33%, RR 2.53, 95% CI 1.9-3.3) than non-VTE patients. Day 0 and day 3 D-dimer levels were higher in patients who had VTE (Δ +1738 ng/ml at day 0 and Δ +890 ng/ml at day 3 between VTE-positive and negative patients). D-dimer was associated with VTE after adjusting for age (day 0: OR 1.38, 95%CI 1.19-1.60, p<0.001; day 3: OR 1.31, 95%CI 1.08-1.56, p=0.004). Platelet and fibrinogen levels were not significantly different.

**Table 1.** Demographics, clinical and laboratory parameters by COVID-19 status and thromboembolism

|  | All patients by COVID status |  |  |  | COVID-positive patients by VTE |  |  |  |
| --- | --- | --- | --- | --- | --- | --- | --- | --- |
|  | N | COVID negative, N = 78<sup>1</sup> | COVID positive, N = 306<sup>1</sup> | p-value<sup>2</sup> | N | No VTE, N = 293<sup>1</sup> | VTE, N = 13<sup>1</sup> | p-value<sup>2</sup> |
| VTE | 384 | 4 (5.1%) | 13 (4.2%) | 0.8 |  |  |  |  |
| Ethnicity | 384 |  |  | <0.001 | 306 |  |  | 0.5 |
| Non-Hispanic |  | 66 (85%) | 140 (46%) |  |  | 133 (45%) | 7 (54%) |  |
| Hispanic |  | 12 (15%) | 166 (54%) |  |  | 160 (55%) | 6 (46%) |  |
| Age | 384 | 66 (57, 76) | 58 (45, 75) | 0.012 | 306 | 58 (45, 75) | 69 (62, 78) | 0.2 |
| Sex | 384 | 39 (50%) | 144 (47%) | 0.6 | 306 | 138 (47%) | 6 (46%) | >0.9 |
| Heart disease | 384 | 23 (29%) | 48 (16%) | 0.005 | 286 | 30 (26, 34) | 25 (24, 28) | 0.017 |
| Kidney disease | 384 | 20 (26%) | 41 (13%) | 0.008 | 306 | 46 (16%) | 2 (15%) | >0.9 |
| Liver disease | 384 | 7 (9.0%) | 15 (4.9%) | 0.2 | 306 | 40 (14%) | 1 (7.7%) | >0.9 |
| Diabetes | 384 | 28 (36%) | 111 (36%) | >0.9 | 306 | 15 (5.1%) | 0 (0%) | >0.9 |
| Lung disease | 384 | 40 (51%) | 66 (22%) | <0.001 | 306 | 64 (22%) | 2 (15%) | 0.7 |
| Severe disease | 384 | 23 (29%) | 109 (36%) | 0.3 | 306 | 98 (33%) | 11 (85%) | <0.001 |
| Platelets (d0) | 380 | 224 (158, 280) | 208 (161, 280) | 0.7 | 302 | 207 (160, 280) | 253 (179, 307) | 0.2 |
| Platelets (d3) | 292 | 195 (161, 243) | 245 (180, 332) | <0.001 | 235 | 244 (180, 323) | 273 (196, 363) | 0.3 |
| Platelets (d7) | 195 | 218 (172, 268) | 312 (220, 406) | <0.001 | 159 | 306 (220, 400) | 338 (307, 504) | 0.2 |
| CRP (d0) | 366 | 22 (9, 67) | 105 (48, 160) | <0.001 | 294 | 95 (47, 157) | 150 (131, 206) | 0.041 |
| CRP (d3) | 228 | 71 (39, 147) | 117 (55, 168) | 0.13 | 201 | 117 (54, 164) | 165 (78, 240) | 0.13 |
| CRP (d7) | 144 | 72 (22, 148) | 117 (43, 164) | 0.3 | 132 | 96 (42, 164) | 147 (78, 148) | 0.6 |
| D-dimer (d0) | 358 | 1,082 (596, 2,288) | 1,072 (684, 1,808) | 0.8 | 290 | 1,026 (655, 1,721) | 2,764 (1,594, 7,473) | <0.001 |
| D-dimer (d3) | 209 | 1,015 (782, 1,660) | 1,283 (801, 2,152) | 0.2 | 189 | 1,224 (787, 2,066) | 2,114 (1,599, 6,582) | 0.005 |
| D-dimer (d7) | 133 | 1,342 (1,087, 2,370) | 1,999 (1,171, 3,472) | 0.2 | 124 | 1,967 (1,032, 3,374) | 3,350 (1,770, 5,634) | 0.08 |
| Fibrinogen (d0) | 141 | 406 (293, 449) | 603 (533, 692) | <0.001 | 127 | 607 (535, 692) | 564 (483, 630) | 0.4 |
| Fibrinogen (d3) | 98 | 662 (537, 802) | 677 (574, 800) | >0.9 | 94 | 671 (574, 801) | 764 (713, 777) | 0.5 |
| Fibrinogen (d7) | 61 | 821 (821, 821) | 732 (634, 834) | 0.4 | 59 | 715 (626, 830) | 807 (790, 923) | 0.2 |

<sup>1</sup> n (%); Median (IQR)
<sup>2</sup> Fisher's exact test; Pearson's Chi-squared test; Wilcoxon rank sum test

### Plasma coagulation protein levels vs. VTE diagnosis in COVID-19

We selected markers from the Olink and Somalogic libraries representing 31 proteins that are involved in coagulation (Table 2, Table S1). COVID-19-positive patients who developed VTE had higher day 0 plasma levels of factors IX and XI, P-selectin, plasmin, activated protein C, and von Willebrand factor and lower levels of ADAMTS13, antithrombin III, factor VII, protein C, and prolylcarboxypeptidase, in at least one assay (Table 2). Notably, P-selectin and von Willebrand factor levels were significantly increased and ADAMTS13 and factor VII were significantly decreased in both assays.

**Table 2.** Coagulation protein levels of COVID-19 patients measured by two proteome assays in day 0 samples.

|  | Somalogic SomaScan |  |  |  | Olink Explore |  |  |  |
| --- | --- | --- | --- | --- | --- | --- | --- | --- |
|  | N | No VTE, N = 293<sup>1</sup> | VTE, N = 13<sup>1</sup> | p-value<sup>2</sup> | N | No VTE, N = 292<sup>1</sup> | VTE, N = 13<sup>1</sup> | p-value<sup>2</sup> |
| ADAMTS13 | 306 | 13.97 (13.60, 14.25) | 13.37 (12.99, 13.99) | **0.016** | 305 | 4.16 (3.92, 4.37) | 3.90 (3.52, 4.14) | **0.026** |
| Alpha-2 antiplasmin | 306 | 12.59 (12.39, 12.72) | 12.40 (11.91, 12.70) | 0.13 | not included in the protein panel |  |  |  |
| Antithrombin III | 306 | 16.56 (16.44, 16.68) | 16.45 (16.26, 16.50) | **0.022** |  |  |  |  |
| CPB2 | 306 | 13.65 (13.49, 13.88) | 13.69 (13.54, 14.04) | 0.4 |  |  |  |  |
| Factor II | 306 | 17.12 (16.99, 17.26) | 17.04 (16.94, 17.11) | 0.13 |  |  |  |  |
| Factor IIa | 306 | 9.92 (9.54, 10.19) | 9.72 (9.45, 9.89) | 0.1 |  |  |  |  |
| Factor III | 306 | 10.52 (10.38, 10.75) | 10.46 (10.24, 10.85) | 0.6 | 305 | 5.00 (4.63, 5.47) | 5.25 (4.69, 5.57) | 0.4 |
| Factor V | 306 | 14.19 (13.95, 14.45) | 14.20 (14.13, 14.49) | 0.6 | not included in the protein panel |  |  |  |
| Factor VII | 306 | 15.62 (15.24, 15.91) | 15.18 (14.85, 15.33) | **<0.001** | 305 | 3.61 (3.28, 3.87) | 3.28 (3.14, 3.48) | **0.029** |
| Factor VIII | 306 | 14.01 (13.74, 14.27) | 13.89 (13.71, 14.51) | 0.6 | not included in the protein panel |  |  |  |
| Factor IX | 306 | 13.88 (13.71, 14.06) | 14.03 (13.85, 14.27) | 0.15 | 305 | 3.61 (3.28, 3.96) | 4.03 (3.62, 4.15) | **0.02** |
| Factor IXab | 306 | 13.05 (12.88, 13.25) | 13.22 (13.02, 13.47) | 0.2 |  |  |  |  |
| Factor X | 306 | 10.51 (10.31, 10.65) | 10.26 (9.84, 10.54) | 0.076 |  |  |  |  |
| Factor Xa | 306 | 10.76 (10.55, 10.92) | 10.64 (10.11, 10.83) | 0.12 |  |  |  |  |
| Factor XI | 306 | 11.50 (11.26, 11.70) | 11.93 (11.59, 11.98) | **0.002** |  |  |  |  |
| Factor XIII | 306 | 12.26 (12.05, 12.48) | 12.00 (11.96, 12.37) | 0.2 |  |  |  |  |
| FGL2 | 306 | 10.48 (10.30, 10.70) | 10.57 (10.42, 10.88) | 0.3 |  |  |  |  |
| Fibrinogen/Ddimer | 306 | 17.03 (16.89, 17.20) | 17.04 (16.91, 17.28) | >0.9 |  |  |  |  |
| KLKB1 | 306 | 15.02 (14.80, 15.25) | 15.03 (14.80, 15.10) | 0.4 |  |  |  |  |
| KNG1 | 306 | 10.55 (10.13, 10.87) | 10.37 (9.81, 10.65) | 0.3 |  |  |  |  |
| P-selectin | 306 | 13.14 (12.75, 13.54) | 13.53 (13.19, 14.44) | **0.02** | 305 | 2.78 (2.21, 3.26) | 3.56 (2.86, 3.81) | **0.005** |
| Plasmin | 306 | 10.48 (10.32, 10.83) | 10.97 (10.52, 11.00) | **0.004** | not included in the protein panel |  |  |  |
| Plasminogen | 306 | 9.19 (8.97, 9.36) | 9.36 (8.84, 9.45) | 0.4 |  |  |  |  |
| Plasminogen Activator Inhibitor I | 306 | 12.21 (11.63, 12.78) | 12.63 (11.79, 12.89) | 0.3 | 305 | 6.43 (5.69, 7.20) | 7.06 (6.35, 7.35) | 0.2 |
| Prolylcarboxypeptidase | 306 | 13.92 (13.58, 14.22) | 13.65 (13.44, 13.92) | **0.045** | 305 | 2.82 (2.52, 3.10) | 2.77 (2.41, 3.07) | 0.5 |
| Protein C | 306 | 15.93 (15.62, 16.18) | 15.58 (15.02, 15.93) | **0.01** | 305 | 3.28 (2.96, 3.49) | 3.09 (2.81, 3.57) | 0.4 |
| Protein C (activated) | 306 | 9.21 (9.07, 9.41) | 9.47 (9.23, 9.79) | **0.018** |  |  |  |  |
| Protein S | 306 | 11.11 (10.93, 11.31) | 11.17 (10.97, 11.22) | >0.9 |  |  |  |  |
| TBXAS1 | 306 | 10.25 (10.09, 10.43) | 10.33 (9.81, 10.38) | 0.6 | not included in the protein panel |  |  |  |
| Thrombomodulin | not included in the protein panel |  |  |  | 305 | 2.88 (2.57, 3.26) | 3.08 (2.76, 3.20) | 0.4 |
| Thrombospondin 1 | 306 | 10.65 (9.78, 11.33) | 11.34 (10.06, 11.84) | 0.12 | not included in the protein panel |  |  |  |
| Tissue Factor Pathway Inhibitor | 306 | 14.67 (14.40, 14.95) | 14.88 (14.53, 15.23) | 0.3 | 305 | 4.53 (4.25, 4.80) | 4.58 (4.52, 5.05) | **0.14** |
| Tissue Factor Pathway Inhibitor 2 | 306 | 9.03 (8.77, 9.28) | 8.81 (8.61, 9.26) | 0.1 | 305 | 4.44 (3.98, 4.84) | 4.35 (3.92, 4.82) | 0.9 |
| tPA | 306 | 12.97 (12.58, 13.39) | 13.04 (12.88, 13.28) | 0.7 | 305 | 4.41 (3.92, 4.89) | 4.80 (4.45, 5.09) | 0.078 |
| uPA | 306 | 11.44 (11.27, 11.66) | 11.38 (11.00, 11.48) | 0.11 | 305 | 3.08 (2.83, 3.34) | 3.02 (2.74, 3.22) | 0.4 |
| von Willebrand Factor | 306 | 14.89 (14.47, 15.31) | 15.42 (15.07, 15.76) | **0.01** | 305 | 5.70 (5.32, 6.11) | 6.03 (5.77, 6.40) | **0.038** |

<sup>1</sup> n (%); Median (IQR)
<sup>2</sup> Fisher's exact test; Pearson's Chi-squared test; Wilcoxon rank sum test

In multivariable linear regression modeling of each coagulation protein separately – and controlling for possible confounding factors: age, gender, ethnicity, comorbidities, and illness severity – we found that VTE was independently associated with higher day 0 levels of factor XI and P-selectin in the Somalogic panel and factor IX and P-selectin in the Olink panel. Factor IX was not significant in the Somalogic panel, while factor XI was not present in the Olink panel. Furthermore, we found independent association of VTE with lower levels of protein C (in the Somalogic panel) and alpha-2-antiplasmin (only present in the Somalogic panel) (Figure 1). In a subset of COVID-19 positive patients who had high clinical suspicion (i.e. high pre-test probability) of PE or DVT, defined as undergoing CTPE or US during their hospitalization (n=82 total, n=13 confirmed VTE), we also found an independent association of elevated P-selectin with VTE (in both assays), factor IX (Olink only) and factor XI (Somalogic only) on day 0 (Figure 1B).

**Figure 1.**
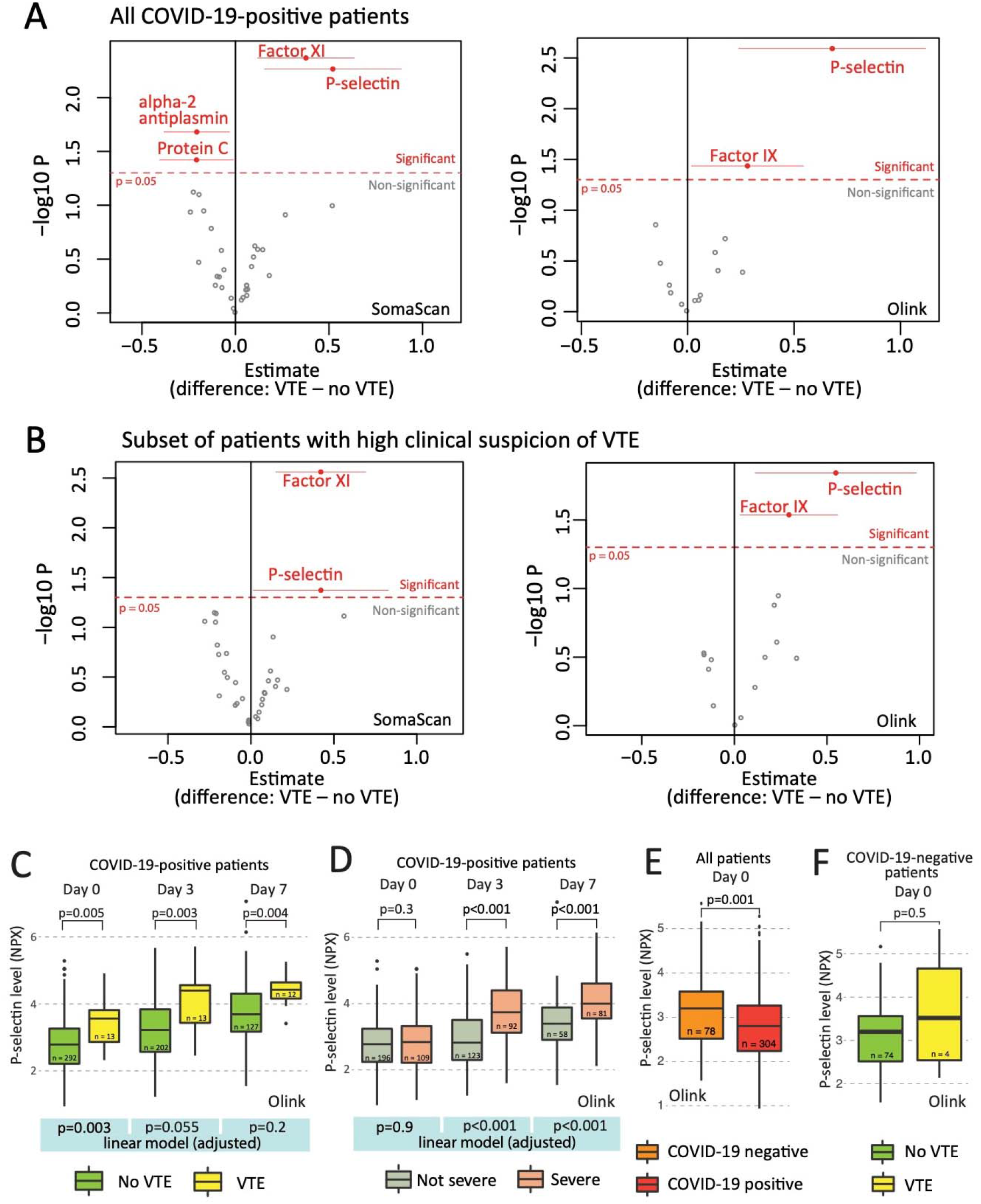
**A** In multivariable linear regression models higher day 0 P-selectin, Factor IX and FXI levels, and lower alpha-2 antiplasmin and Protein C levels were associated with VTE after adjusting for age, sex, ethnicity, comorbidities, and severity of COVID-19. Red color marks proteins with significant difference (p<0.05). Horizontal lines for significant proteins represent confidence intervals (95%). **B** In a subset of N=82 confirmed COVID-19 positive patients who underwent CTPE or ultrasound because of clinical suspicion of PE or DVT (N=13 confirmed VTE cases) an association of P-selectin level with VTE risk was confirmed. **C** Temporal dynamics of plasma P-selectin level in COVID-positive patients (Olink assay). **D** Temporal dynamics of P-selectin in severe vs not severe patients **E** COVID-19 positive versus negative patients. **F** P-selectin level in VTE vs. no VTE COVID-19 negative patients. In **C-F**, univariate tests were non-parametric Kruskal-Wallis tests. Multivariable linear models were adjusted as in Panel A.

Given the independent significance of P-selectin in both assays, we further analyzed its temporal dynamics. Using the Olink data we observed a steady increase in the plasma concentration of P-selectin in VTE patients over time. Median plasma level of P-selectin in VTE-positive patients was significantly higher than in VTE-negative patients on each day (Figure 1C). In adjusted linear regression models, P-selectin on day 0 was independently associated with VTE, which progressively diminished on days 3 and 7 (Figure 1C). The diminished association between P- selectin and VTE was accompanied by a strong independent association between illness severity and VTE on days 3 and 7 that was not present on day 0 (Figure 1D). Thus, illness severity may increasingly confound the relationship between P-selectin and VTE over time.

Figure 1Despite this confounding, P-selectin levels were associated with VTE at all time points. But our findings suggest that P-selectin might be of particular value early in the disease course, prior to any influence of illness severity on P-selectin levels, and at a time when the degree of severity may still be unclear clinically. Thus, P-selectin is a promising candidate for a predictive biomarker of VTE upon hospital presentation.

We also tested whether P-selectin levels were related to COVID-19 status and found that the median level was higher in the COVID-negative cohort (Figure 1E). Among the COVID-19 negative patients, although our data show higher median P-selectin values for VTE patients, this difference was not significant; however, with only 4 VTE events in 78 COVID-19 negative patients, there was little power to demonstrate significance (Figure 1F). Of note, age and comorbidities including chronic kidney disease were more prevalent in the COVID-19 negative cohort, which may be contributory to higher P-selectin levels in this group.

To evaluate the predictive performance of P-selectin in COVID-19-positive patients in the context of clinical biomarkers associated with VTE and illness severity, we created logistic regression models using P-selectin, D-dimer and fibrinogen as independent variables and analyzed the receiver operator characteristic (ROC) curves. We found that although the area under the ROC curve (AUC) of the P-selectin model was lower than that of D-dimer, the combined P-selectin + D-dimer model performed better than D-dimer alone (AUC 0.834 vs. 0.783, Figure 2). Both the fibrinogen and fibrinogen + P-selectin models had lower AUCs than the P-selectin model alone.

**Figure 2.**
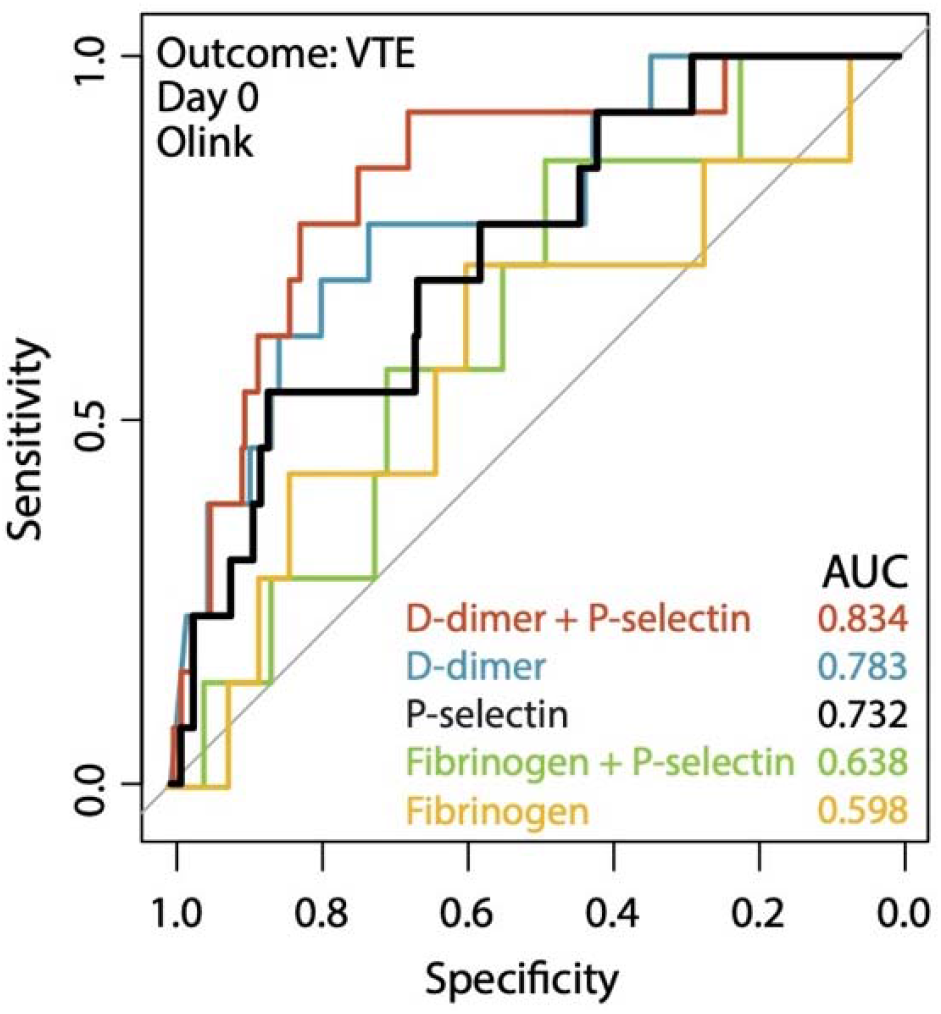
Predictive performance of P-selectin, D-dimer and Fibrinogen for VTE. Receiver operator characteristic curves were plotted for logistic regression models that take Day 0 Olink protein levels of P-selectin (black) and D-dimer (blue), Fibrinogen (yellow) and their combinations (red and green) as predictors and venous thromboembolic events as outcome.

## Discussion

Our study demonstrates a consistent association between serum P-selectin levels and the development of VTE in a large cohort of admitted patients with COVID-19 across two independent proteomic platforms. P-selectin was associated with VTE upon presentation to the hospital after adjusting for age, comorbidities, and illness severity, implicating endothelial activation in the development of VTE and demonstrating promise for P-selectin as an early biomarker in predicting presence or development of VTE in COVID-19. P-selectin added to the value of D-dimer alone in predicting VTE diagnosis during hospitalization.

Thromboembolic events are a known complication of COVID-19 (Malas et al., 2020). Whereas several mechanisms have been implicated in hemostatic imbalance, a clear understanding of the pathogenesis and factors that may lead to or prevent COVID-19 associated coagulopathy (CAC) is lacking. It has been suggested that immunothrombosis, neutrophil extracellular trap formation (NET-osis), and endothelial and platelet activation/damage are involved (Bongiovanni et al., 2021; Goshua et al., 2020; Hottz et al., 2020; Zhang et al., 2020). Laboratory and histopathological findings suggest that features of COVID-19 associated coagulopathy are unique and are distinct from those of disseminated intravascular coagulation or thrombotic microangiopathies (Iba et al., 2020; Martín-Rojas et al., 2020).

Prior studies have shown that elevated D-dimer, fibrinogen, and prolonged prothrombin time characterize COVID-19-associated coagulopathy and thromboembolism, and are also associated with ICU-level care and death (Tang et al., 2020). Other coagulation biomarkers not typically used in acute clinical care, such as factors V, VII, VIII, X and ADAMTS13, von Willebrand factor, thrombomodulin, protein C, and prothrombin have been associated with varying clinical outcomes in COVID-19 (Goshua et al., 2020; Ladikou et al., 2020; Martín-Rojas et al., 2020; Stefely et al., 2020; Tiscia et al., 2020). Elevated factor V levels have been linked specifically to VTE in severe COVID-19 (Stefely et al., 2020). Although we found that the levels of a number of these proteins were increased (namely von Willebrand factor, protein C) or decreased (ADAMTS13, factor VII) in the day 0 serum of patients who developed VTE in our cohort, none of them were significantly associated with VTE when adjusting for confounders. This was similar in the case of other assayed proteins that have not been previously reported in COVID-19 (antithrombin III and prolylcarboxypeptidase). Factors IX and XI were significantly associated with VTE also in the adjusted models, but only in one platform (Olink and Somalogic, respectively).

Our analysis demonstrated that P-selectin had a prominent association with the development of VTE in hospitalized COVID-19 patients, and in particular this association was apparent upon hospital presentation, independent of disease severity and other relevant confounders. P-selectin is an integral membrane protein of activated platelets and endothelial cells that mediates leukocyte rolling (adhesion to neutrophils and monocytes). Aside from participating in the inflammatory response, P-selectin contributes to immunothrombosis by facilitating formation of endothelium-leukocyte and platelet-leukocyte aggregates and NET-osis (Etulain et al., 2015; Zucoloto & Jenne, 2019). P-selectin is shed into the circulation and increased plasma levels are considered a marker of endothelial cell and platelet activation and damage (Goshua et al., 2020).

Importantly, a possible pathogenetic role of P-selectin in ARDS and more specifically coronavirus-induced ARDS has been suggested (Neri et al., 2020), and prior studies have demonstrated a correlation between P-selectin levels and disease severity (Barrett et al., 2020; Goshua et al., 2020). Increased platelet surface expression of P-selectin in COVID-19 has also been shown in plasma-based cytometry studies (Bongiovanni et al., 2021; Zhang et al., 2020). However, an association between plasma P-selectin levels and thrombosis in COVID-19 has not been previously reported.

Our findings support the use of P-selectin as a candidate early biomarker for risk stratification of VTE in COVID-19. Although not superior to D-dimer in its ability to discriminate VTE events when analyzed separately, P-selectin increases the discriminatory ability when used in combination with D-dimer, compared with D-dimer alone. Consistent with prior literature, our data show an association between plasma P-selectin levels and disease severity, although only on hospital days 3 and 7, and not on day 0. This suggests that P-selectin may be a late marker of severity and is evidence of a potentially delayed role of endothelial activity or injury in severe COVID-19. In contrast, D-dimer is strongly associated with severity in this cohort starting on day 0 (Filbin et al., 2021), suggesting acute inflammation early in severe disease, measurable by this acute phase reactant, early in severe disease.

The link between P-selectin and VTE risk provides further support for the evaluation of P-selectin blockade in the prevention or treatment of COVID-19-associated thrombosis and embolism (Neri et al., 2020). By preventing adherence of leucocytes and platelets to the vessel wall, P-selectin blockade can potentially decrease inflammation and thrombosis (Lowenstein & Solomon, 2020). Two monoclonal antibodies against P-selectin have been developed to prevent vaso-occlusive crisis in sickle cell disease (Ataga et al., 2017). Currently, there is one Phase 2 clinical trial investigating crizanlizumab in COVID-19 vasculopathy (NCT04435184). The other anti-P-selectin antibody, inclacumab, has not yet been studied in COVID-19.

We found plasma P-selectin levels to be generally lower in COVID-19-positive compared to COVID-19-negative patients. A possible explanation is the higher proportion of older patients and those with pre-existing conditions, in particular kidney disease in the COVID19-negative cohort (Table 1). Therefore, a clinical threshold for P-selectin levels to guide diagnostic imaging for VTE might be optimized by age or renal function adjustment, similar to age-adjusted clinical rules for d-dimer and predicting VTE risk (Righini et al., 2014). The optimal application of P-selectin in the clinical setting needs to be further clarified with larger and independent cohorts that employ methods to measure absolute plasma concentrations (e.g. ELISA).

In this post-hoc analysis of proteomic data from a prospective study, we simultaneously analyzed the plasma level of 31 proteins. Compared with prior reports on this topic, the strengths of our study are that we 1) obtained serial blood samples on fixed days starting at Emergency Department presentation, 2) enrolled all patients that presented to the Emergency Department with respiratory distress prior to knowledge of COVID-19 status, and thus obtained a significantly ill COVID-negative control group, 3) had the power to perform adjusted analysis for covariates, and 4) enrolled a large sample size with high density of severe illness. The symptomatic control group had a similar distribution of severe and non-severe patients that allowed a comparison of COVID-19-positive and negative patients.

Our study has limitations. First, the single-center observational study design necessitates external validation. Second, a causal link between endothelial activation, P-selectin levels, and VTE cannot be made. Third, serial sampling was only done in those still hospitalized, which biases later samples toward sicker patients. Fourth, the biomarkers were limited to those available on the two proteomic platforms. Fifth, both proteomic platforms provide relative protein abundances and not absolute plasma concentrations. Lastly, the relative contribution of various tissues to the plasma proteome is not knowable.

In this study, we evaluated the possible association of a comprehensive set of coagulation-related proteins with VTE in COVID-19, showing that P-selectin is independently associated with development of VTE. This supports the mechanistic role of platelet/endothelial-activation in COVID-19-related thromboembolism. We found that measuring P-selectin levels at day 0 of hospitalization increases the diagnostic value of D-dimer alone, and that a combination of the two biomarkers (D-dimer and P-selectin) may provide a more accurate prediction of VTE in the setting of COVID-19. Further studies to address the utility of P-selectin as a predictive biomarker of VTE are warranted.

## Methods

### Cohort and sampling, and clinical data

A total of 384 patients were enrolled in this study, who presented with respiratory distress to the Emergency Department of an urban, academic hospital in Boston, MA between 3/24/2020 and 4/30/2020 during the first peak of COVID-19 surge, as described previously (Filbin et al., 2021). Inclusion criteria were age ≥18 years, clinical concern for COVID-19, and acute respiratory distress with at least one of the following: 1) tachypnea (≥22 breaths/min), 2) oxygen saturation ≤92% on room air, 3) need for supplemental oxygen, or 4) positive-pressure ventilation. The institutional review board waived informed consent. Of the 384 enrolled, 306 patients were confirmed positive for SARS-CoV-2 by reverse transcriptase polymerase chain reaction (RT-PCR).; 78 patients were used as symptomatic COVID-19 negative controls.

Specimen collection and banking was performed as previously described, (Filbin et al., 2021). Samples were obtained on the day of admission (day 0) from all patients (N=384), and subsequently on day 3 (N=217) and 7 (N=143) from COVID-19-positive patients still hospitalized, yielding a total of 744 samples. Clinical course was followed to 28 days post-enrollment or until hospital discharge if that occurred after 28 days. Clinical data was obtained from electronic health records. Patients who were intubated and/or died at any time during the 28-day follow-up period were classified as *severe*.

### Biomarker assay and selection

Plasma biomarkers were measured using the Olink Explore 1536 Proximity Extension Assay and the Somalogic SomaScan Platform^®^ aptamer assay, as described previously (Filbin et al., 2021). Data from both assays were analyzed after normalization and log_2_-transformation. Coagulation biomarkers were selected *a priori* from the Olink and Somalogic protein libraries based on literature and hypothesized relevance. The selected markers included pro- and anticoagulant proteins in both their inactivated and activated forms, when available.

### Analysis

All statistical analysis was done in RStudio (version 4.0.3). Plots were generated using standard functions and the *ggplot2* package in R. Group comparisons were done with the *tbl_summary()* function. Multivariable linear regression models were fit independently to each protein using the *lm* function in R, using protein values as the dependent variables. The multivariable models adjusted for age, gender, ethnicity, heart disease, hypertension, lung disease, kidney disease, liver disease, diabetes, and illness severity. Logistic regression models were created using the *lm* function and ROC curves were plotted using the *pROC* package. Code is available: https://github.com/bank-fenyves/COVIDprot

## Data Availability

The data that supports the findings of this study are available in the supplementary material of this article.

https://github.com/bank-fenyves/COVIDprot

## Author Contributions

Conceptualization: BGF, AM, NH, MRF, MBG.

Resources: MRF, NH, MBG, IG.

Methodology: BGF, AM, NH, MRF, MBG.

Investigation: All investigators.

Formal Analysis: BGF.

Writing – Original Draft: BGF.

Writing – Review & Editing: BGF, AM, NH, MRF, MBG.

## Funding and disclosures

Direct funding for this project was provided in part by a grant from the National Institute of Health (N.H., U19 AI082630), an American Lung Association COVID-19 Action Initiative grant (M.B.G.), and grants from the Executive Committee on Research at MGH (M.B.G.). We thank Arthur, Sandra and Sarah Irving for a gift that enabled this study and funded the David P. Ryan, MD Endowed Chair in Cancer Research (N.H.). B.G.F was supported by the Rosztoczy Foundation Scholarship, by the Semmelweis University (EFOP-3.6.3-VEKOP-16-2017-00009) and by the Human Capacities Grant Management Office in Hungary (Emberi Eroforrasok Miniszteriuma, NTP-NFTÖ-20-B-0076). This work was also supported by the Harvard Catalyst/Harvard Clinical and Translational Science Center (National Center for Advancing Translational Sciences, National Institutes of Health, UL1 TR 002541-01). We are very grateful for the generous contributions of Olink Proteomics Inc. and Somalogic Inc. for all in this work, without which our findings would not have been possible.

In addition, the authors declare the following interests:

AM – NIH / NCI T32 2T32CA071345-21A1 (unrelated to this work). A.M. is a consultant for Third Rock Ventures, Abata Therapeutics, Asher Biotherapeutics and Rheos Medicines, and holds equity in Abata Therapeutics and Asher Biotherapeutics (all unrelated to this work). N.H. holds equity in BioNTech and is a consultant for Related Sciences. MRF – Research grants received from Day Zero Diagnostics, Rapid Pathogen Screening Inc., Nihon Kohden Corporation (all unrelated to this work).

## Ethics approval statement

All study procedures involving human subjects were approved by the Mass General Brigham (formerly Partners) Human Research Committee, the governing institutional review board at Massachusetts General Hospital.

## Patient consent statement

A waiver of informed consent was approved in compliance with the Code of Federal Regulations (45CFR 46, 2018 Common Rule).

## Permission to reproduce material from other sources

Not applicable.

## Clinical trial registration

Not applicable.

